# Human CA1 and subiculum activity forecast stroke chronicity

**DOI:** 10.1101/2020.01.19.20017996

**Authors:** Diogo Santos-Pata, Belén Rubio Ballester, Riccardo Zucca, Carlos Alberto Stefano Filho, Sara Regina Almeida, Li Li Min, Gabriela Castellano, Paul FMJ Verschure

## Abstract

Following a stroke, the brain undergoes a process of neuronal reorganization to compensate for structural damage and cope with functionality loss. Increases in stroke-induced neurogenesis rates in the dentate gyrus and neural migration from the hippocampus towards the affected site have been observed, suggesting that the hippocampus is involved in functionality gains and neural reorganization. Despite the observed hippocampal contributions to structural changes, the hippocampal physiology for stroke recovery has been poorly characterized. To this end, we measured resting-state whole-brain activity from non-hippocampal stroke survivors (n=13) during functional MRI scanning. Analysis of multiple hippocampal subregions revealed that the voxel activity of hippocampal readout sites (CA1 and subiculum) forecast the patient’s chronicity stage stronger than early regions of the hippocampal circuit. Furthermore, we observed hemispheric-specific contributions to chronicity forecasting, raising the hypothesis that left and right hippocampus are functionally dissociable during recovery. In addition, we suggest that in contrast with whole-brain analysis, the monitoring of segregated and specialized sub-networks after stroke potentially reveals detailed aspects of stroke recovery. Altogether, our results shed light on the contribution of the subcortical-cortical interplay for neural reorganization and highlight new avenues for stroke rehabilitation.

## Introduction

The occlusion or rupture of cerebral vessels leads to the immediate deprivation of metabolic substrate to brain cells, causing a cascade of excitotoxicity, neural death, and functional loss^1^. A few minutes after the lesion, structural damage is already visible and impairment appears. Besides the damage surrounding the lesion, the resulting axonal loss leads to the deafferentation of distal neural units, resulting in a global network breakdown, the so-called ‘diaschisis’, through the hypoperfusion and hypometabolism of focal brain areas that are remote but directly connected to the injured site^2^. The injury triggers a structured torrent of processes enhancing neuroplasticity and resilience^3^. The temporal structure and interaction of these processes remain unknown. However, a large body of evidence from animal models suggests that they may combine neurogenesis, gliogenesis, axonal sprouting, and the rebalancing of excitation and inhibition in cortical networks^4-7^.

In humans, imaging and behavioral studies have shown that the brain of an acute stroke patient presents a remarkable potential for plastic changes^8,9^ to restore behaviour to near-normal function. During the first days following the stroke, as the dormant cells in the ischemic penumbra oxygenate and acquire functionality, the patient experiences fast recovery, a process often called “spontaneous recovery”^10^. However, three to four weeks after the stroke, this extraordinary initial capacity for recovery seems to vanish^11^. Altogether, results from randomized controlled trials elucidate that the process of neurological repair may follow two general principles: proportionality and temporal structure. The former grounds on the observation that recovery is proportional to the initial functional loss; in other words, those patients who display a more substantial impairment due to the injury do experience more significant recovery. The temporal structure principle refers to a generally heightened responsiveness to treatment at the more acute stage and links to the notion of a sensitive period for recovery. The mechanisms responsible for each of these principles remain unclear. However, recent studies have investigated interventions that could interfere with the recovery process, either by rising recovery rates or by re-opening the sensitive period for recovery at the chronic stage^9^. A recent study with mice has shown that the induction of a second stroke at the chronic stage re-opens a sensitive period and - if combined with training - it mediates full recovery of those impairments that derived from the first stroke^12^. Thus, the observations from^12^ emphasize the close interaction between behaviour and those enhanced plasticity mechanisms that are unique to the injured brain.

Aiming at inducing and promoting stroke-derived neuroplasticity, behavioural, and pharmacological treatments are targeting the expression of growth factors. The pharmacological treatment with selective serotonin reuptake inhibitors (e.g., fluoxetine) enhances the expression of brain-derived neurotrophic factor (BDNF)^13–15^, a protein related to canonical nerve growth factor, which promotes resilience and facilitates neural reorganization. Following this line of research, the administration of fluoxetine has shown to increase the BDNF gene expression, to prolong critical periods of recovery in animal stroke models^16^ and to facilitate motor recovery in humans^17^. Intracerebroventricular and intravenous infusion of BDNF increases the number of pyramidal cells in CA1, reduces the infarct volume, and promotes motor improvements^18^. The effects of physical exercise seem analogous to the effects of selective serotonin reuptake inhibitors, and rodent models support that physical exercise increases the hippocampal levels of BDNF^19,20^.

Similarly, the overexpression of fibroblast growth factor (FGF-2) has been shown to promote hippocampal neurogenesis after stroke^21^. More specifically, two main structures - the subgranular zone of the hippocampal dentate gyrus and the subventricular zone - are the primary substrate of neural stem cells in the adult brain^22–24^. Recent studies have shown hippocampal stroke-induced neurogenesis in the human cerebral cortex, suggesting the critical contribution of the hippocampus to neural reorganization and functional restoration^25,26^.

Further to its contribution to recovery via differentiation of stem cells, additional hippocampal structural biomarkers associated with post-stroke reorganization have been identified. Werden and colleagues^27^ have found smaller hippocampal volumes and extensive white matter hyperintensity in the acute phase of first-ever non-demented stroke patients compared to healthy controls, which might be due to reductions in cerebral blood flow^28^. At the chronic stage (3 months), the contralesional hippocampus generally shows a 4% volume loss from the hyperacute stage (2 hours after stroke)^29^. This loss has been shown to correlate with a decline in memory performance^30^.

A number of rodent and human studies investigating the functional role of hippocampal subregions support their distinct contributions to cognitive aspects such as spatial representations^31, 32^, pattern separation in the dentate gyrus^33^, pattern completion in CA3^34^, temporal ordering in CA1^35^, memory retrieval in subiculum^36,37^ and spatial decision-making^38,39^.

Altogether, this set of evidence supports the hypothesis that non-focal or connectional diaschisis due to stroke may lead to functional and structural alterations to the hippocampal complex, however, despite the evidence supporting the role of the hippocampus in brain repair, few studies have quantified the physiological integrity of hippocampal subregions post stroke. Here, we conduct an exploratory analysis on the resting-state fMRI of non-hippocampal stroke patients to assess the specific changes in hippocampal subregions. We observe that the activity of hippocampal readout sites CA1 and subiculum correlates with the patient’s chronicity. Furthermore, we observed that this correlation is hemispheric-specific, raising the hypothesis that left and right hippocampus are functionally dissociable during recovery.

**Table 1.** Distribution of patient characteristics grouped by chronicity

| Chronicity | TSS (days) | N | Female | Stroke left | Age (mean/std) |
| --- | --- | --- | --- | --- | --- |
| Acute | 30 | 11 | 4 | 5 | 64.45±9.76 |
| Subacute | 90 | 12 | 4 | 5 | 64.0±8.96 |
| Chronic | 180 | 8 | 1 | 3 | 62.62±9.71 |

### 1 Results and Discussion

Correlates of functional gains and body structure recovery, the behavioural counterpart of neural repair, were measured through standard clinical scales. The National Institutes of Health Stroke Scale (NIHSS)^40^ was used to assess stroke severity in patients at acute (<1 month), subacute (1-3 months) and chronic (>6 months) stages after stroke. Decreases in NIHSS scoring, reflecting recovery, were observed depending on the chronicity stage of the patient (acute 5 ± 4.06, subacute 3.3 ± 2.3, chronic 2.5 ± 2.9, mean ± std). Performance-based improvements in recovery were further confirmed by an increase in the Fugl-Meyer motor balance assessment scale^41^ (acute 9.2 ± 2.6, subacute 9.5 ± 2.2, chronic 11.25 ± 2.8, mean ± std), as well as in the Total EDT(23) scale (acute 16.4 ± 4.2, subacute 17.5 ± 4.3, chronic 18.25 ± 4.7, mean ± std) (figure 1).

**Figure 1.**
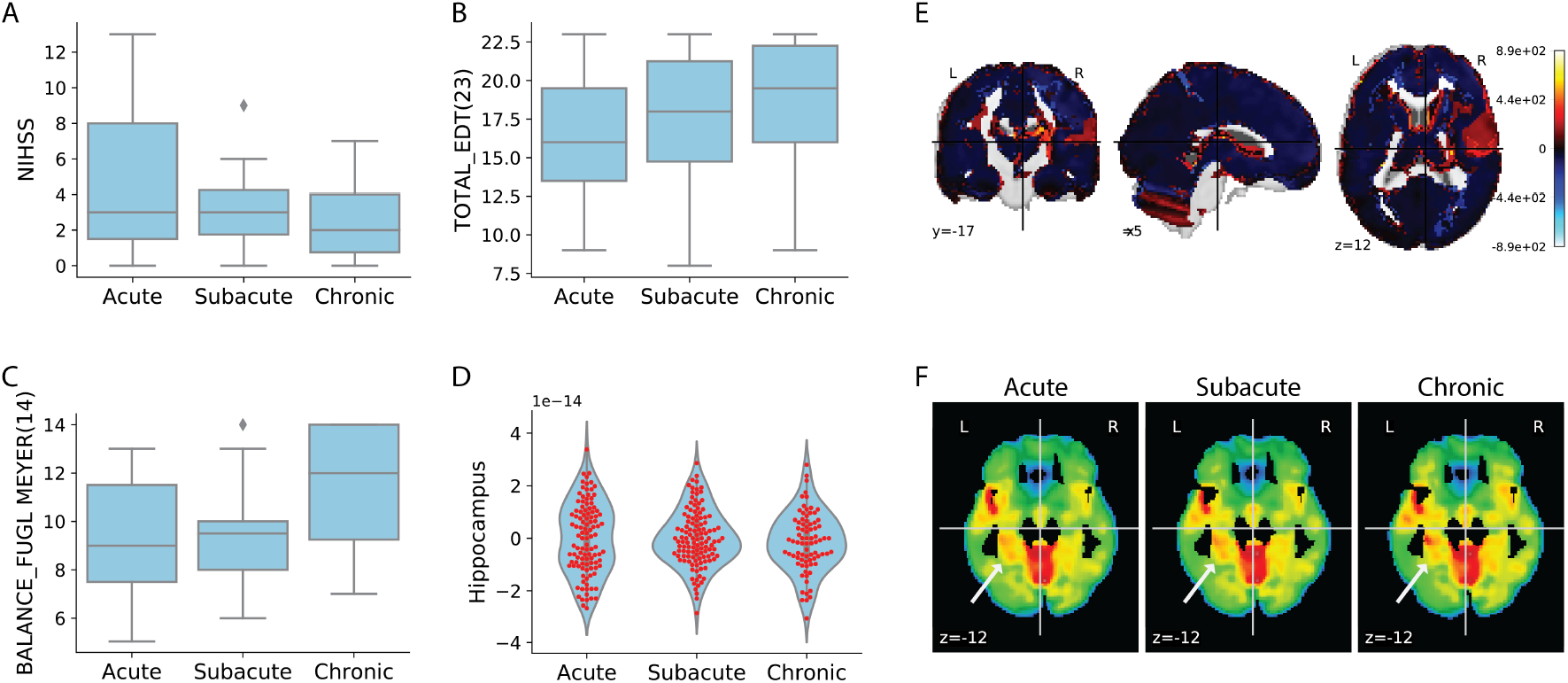
Functional recovery and hippocampal characterization. **A-C** Distribution of clinical assessment scoring per chronicity stage. **A** NIHSS, acute 5 ± 4.06, subacute 3.3 ± 2.3, chronic 2.5 ± 2.9, mean ± std. **B** Total EDT, acute 16.4 ± 4.2, subacute 17.5 ± 4.3, chronic 18.25 ± 4.7, mean ± std. **C** Fugl-Meyer motor balance, acute 9.2 ± 2.6, subacute 9.5 ± 2.2, chronic 11.25 ± 2.8, mean ± std. **D** Overall hippocampal activity is sustained during recovery. Acute −5.47 × 10^−17^±1.34 × 10^−14^; Subacute 1.10 × 10^−16^ ± 1.07 × 10^−14^; Chronic −1.47 × 10^−15^ ± 1.12 × 10^−14^, mean ± std. **E** Across subject contrast between acute and chronic phases. **F** Representative patient showing higher activation in the left medial temporal lobe section at the chronic stage.

Neither overall or hemispheric-specific fMRI hippocampi activity differed across chronicity stages (figure 1-D and figure 2). Nevertheless, within-subject analysis of longitudinal data (n=6) revealed higher resting-state activity in the right temporal lobe at the acute compared to chronic stages (figure 1-E). On the contrary, through visual inspection, we observed punctuated increased activity at later stages of recovery in the contralesional hippocampal complex (see figure 1-F for a representative patient). Neither the dentate gyrus, one of the main sites for neurogenesis, nor other early-hippocampal subregions revealed to have their activity modulated depending on chronicity.

**Figure 2.**
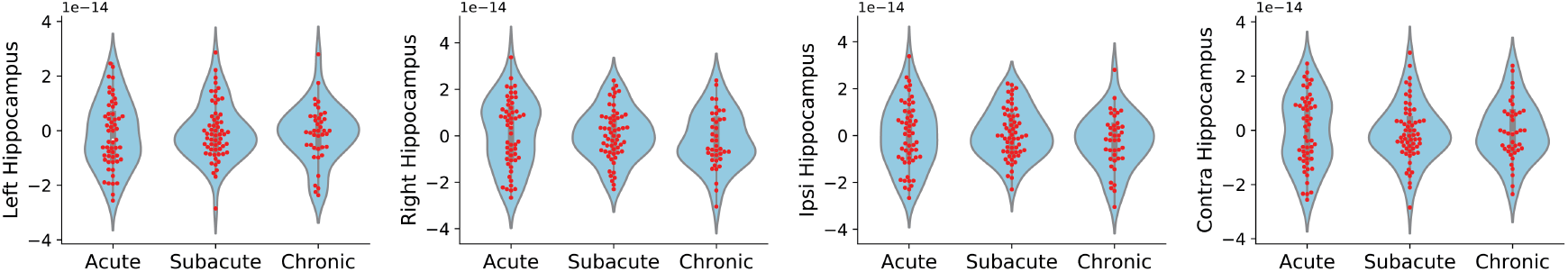
Whole-hippocampus activity is not modulated by chronicity stages. **E** BOLD activity grouped by chronicity stages for left, right, ipsilateral, and contralateral hippocampus.

Interestingly, we found a significant decrease in the ipsilateral CA1 voxel activity from subacute to the chronic stage of recovery (p<0.05, Mann-Whitney U test, figure 3-A), and a significant increase in the contralateral CA1 voxel activity between those stages (p<0.05, Mann-Whitney U test, figure 3-A). Similarly, we found a positive trend in activity for left subicular voxels from early to late chronic stages, with a significant difference between acute and chronic stages (p<0.05, Mann-Whitney U test, figure 3-B). Moreover, a negative trend was found for the right subiculum where voxels’ activity at the chronic stage were significantly lower when compared with both acute (p<0.01, Mann-Whitney U test, figure 3-B) and subacute (p<0.05, Mann-Whitney U test, figure 3-B) stages.

Overall, our results support that cerebral accidents trigger brain-wise global remote neurophysiological changes, leading to changes in the strength and direction of neural pathways and connectivity between cortical and subcortical brain areas. Our investigation on the physiological response of the hippocampus after stroke reveals that the hippocampal output regions, namely CA1 and subiculum, reflect changes in BOLD activity specific to the patient’s chronicity, suggesting that CA1 engages intra-hemisphere networks after cerebral infarcts. Furthermore, as the CA1 region is involved in temporal pattern separation, temporal ordering and memory formation^35,37,42^, and subicular zones are needed for hippocampal-mediated memory retrieval^36,37^, changes in BOLD activation at the late chronic stage suggest the functional reintegration of such subregions in the recovery of mnemonic processes. We did not observe significant differences in the dentate gyrus, as it would be expected in the case on enhanced neurogenesis and an associated increase of astrocytes activity after injury. One possibility for the unseen changes in chronicity-specific dentate gyrus activity might be due to the method used here. As newborn cells develop in the dentate gyrus, they are recruited and migrate towards the damaged zone. Thus, such a transient period is unlikely to be captured during the short scanning sessions. Our findings highlight the global impact of focal lesions on the whole brain network architecture. These results support a holistic approach towards functional recovery, suggesting that effective neurorehabilitation protocols for stroke survivors should combine motor and cognitive training.

**Figure 3.**
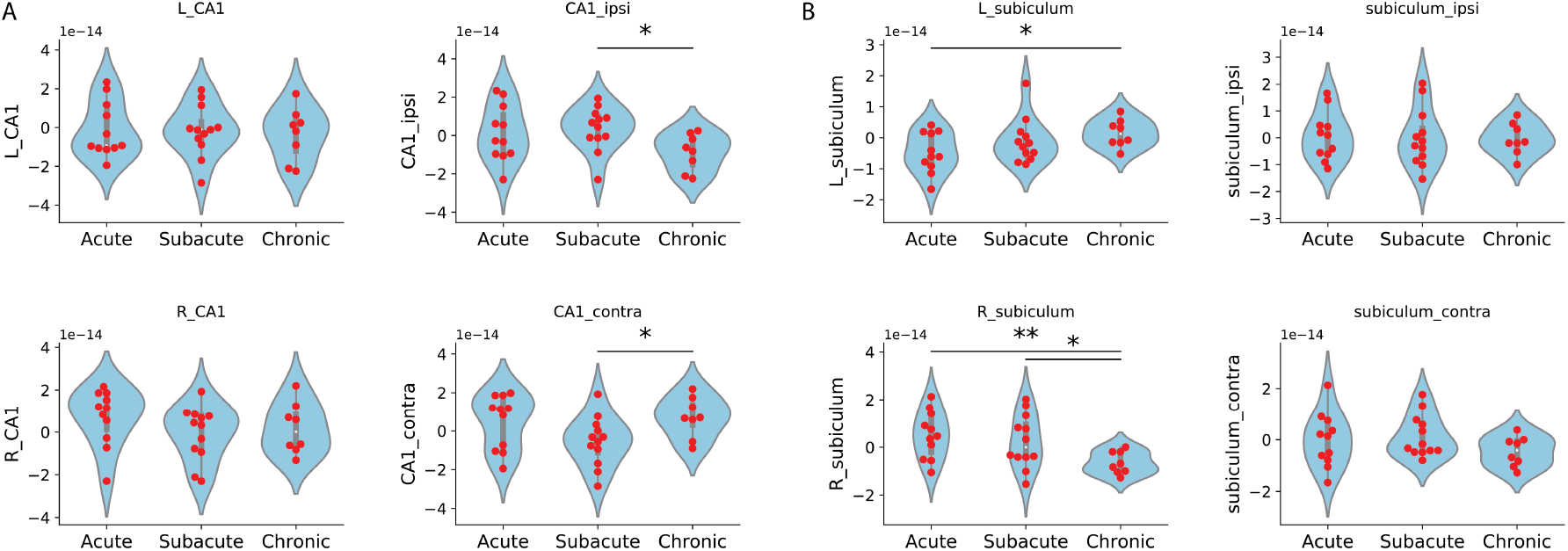
CA1 and Subiculum activity split. **A** CA1 lesional (but not hemispheric) split reveals activity modulation between subacute and chronic periods with decreases in ipsilateral (Mannwhitney-u test, stat=19, p=0.01) and increases in the contralateral sites (Mannwhitney-u test, stat=23, p=0.02). **B** Subiculum hemispheric site split reveals activity modulation along the chronicity stages with increased activity from the actute to chronic stage in the left (Mannwhitney-u test, stat=19, p=0.02) and right (Mannwhitney-u test, stat=13, p=0.006) subiculum. Shorter, but significant increases were also observed from the subacute to the chronic stage in the right subiculum (Mannwhitney-u test, stat=12, p=0.04).

#### Supporting Information (SI)

##### SI Datasets

The dataset used to generate all the quantification figures in this article is supplied in the supplementary section. This file is published in raw format and will not be edited or composed.

### 2 Methods

Thirteen stroke patients (5 female, age 64.2 ±9.0) from the Clinics Hospital of University of Campinas, Brazil, met inclusion criteria and gave consent to participate in this study. The inclusion criteria required that the participant’s stroke onset was at months 1 (acute), 3 (subacute), and/or 6 (chronic) prior to the scanning sessions and that patients achieved mild, but not severe, impairment scoring on the three standardized clinical scales used for evaluation (NIHSS, Fugl-Meyer, EDT(23)).

Each experimental session consisted of 6 minutes of fMRI scanning during resting. After preprocessing, we discarded the first five volumes for each scanning session allowing for T1 equilibration effects, and segmented and grouped voxels belonging to distinct hippocampal subregions defined by the hippocampal fMRI atlas.

Next, we (mean) averaged the overall activity of individual subregions collapsing their dynamics in the time domain. All statistical testing was performed using the subregions averaged activity and grouped based on the chronicity stage.

#### 2.1 fMRI data acquisition and preprocessing

Resting-state blood oxygenation level-dependent (BOLD) signals were recorded using a 3.0T magnetic resonance imaging system (Achieva, Philips HealthCare, Best, The Netherlands) at acute (1 month), subacute (3 months) and chronic (6 months) stages after stroke. Besides, a functional and neurophysiological screening was carried out for each of the subjects. The MRI dataset included one T1-weighted anatomical image (isotropic voxel of 1 mm^3^, repetitions time (TR) of 7 ms, echo time (TE) of 3.2 ms, image matrix of 240×240×180, and one T2*-weighted functional image (voxel size = 3×3×3.6 mm^3^, gap of 0.6 mm, TR of 2 s, TE of 30 ms, image matrix = 80×80×40, 180 volumes). The fMRI images were acquired with an echo-planar imaging (EPI) protocol, isotropic voxels of 3 mm each, 39 slices, a repetition time of 2 s, echo time of 30 ms, a flip-angle of 90°, 180-time points and a field-of-view of 240×240×117. Preprocessing of the functional images was done with the UF2C package in Matlab (^43^, https://www.lniunicamp.com/uf2c). Images were reoriented to set the anterior commissure as the origin of the space and realigned to correct for head movements using rigid body transformations. Functional images were coregistered to the structural ones, and the latter was segmented into white matter, gray matter, and cerebrospinal fluid. All images were then normalized to standard space, using the Montreal Neurological Institute (MNI) template^44^. Additionally, functional images were smoothed with a Gaussian kernel (FWHM 6×6×6 mm^3^) and linearly detrended. Movement parameters and white matter and cerebrospinal fluid signals were regressed out from the BOLD signal. Finally, time series were band-pass filtered to obtain the low resting state frequencies of interest between 0.008 and 0.1 Hz. Hippocampal regions of interest were then selected accordingly to^45,46^ and hippocampal segmentation based on the atlas from (http://cobralab.ca/atlases/Hippocampus-subfields/). After subregion identification, we averaged each field-voxel activity from the resting state acquisition time series, resulting in one activity-scalar per hippocampal subregion from each acquisition session. Thereafter, we performed group-analysis with splitting based on the patient’s chronicity level at the time of the acquisition. Approval for this study was obtained from the Ethics Committee of the University of Campinas.

## Data Availability

Data will be made public upon article acceptance.

## Acknowledgements

We thank all the patients who participated in this study. This project was funded by the European Research Council under grant agreement 286341196 (CDAC), the EIT Health, supported by EIT, a body of the European Union, and the Virtual Brain Cloud (VBC) H2020-EU.3.1.5.3., ID 826421, Research and Innovation action.

